# The Public Health Impact of Delaying a Second Dose of the BNT162b2 or mRNA-1273 COVID-19 Vaccine

**DOI:** 10.1101/2021.02.23.21252299

**Authors:** Santiago Romero-Brufau, Ayush Chopra, Alex J Ryu, Esma Gel, Ramesh Raskar, Walter Kremers, Karen Anderson, Jayakumar Subramanian, Balaji Krishnamurthy, Abhishek Singh, Kalyan Pasupathy, Yue Dong, John C O’Horo, Walter R Wilson, Oscar Mitchell, Thomas C Kingsley

## Abstract

**Objectives:** To estimate population health outcomes under delayedsecond dose versus standard schedule SARS-CoV-2 mRNA vaccination.

**Design:** Agent-based modeling on a simulated population of 100,000 based on a real-world US county. The simulation runs were replicated 10 times. To test the robustness of these findings, simulations were performed under different estimates for single-dose efficacy and vaccine administration rates, and under the possibility that a vaccine prevents only symptoms but not asymptomatic spread.

**Setting:** population level simulation.

**Participants:** 100,000 agents are included in the simulation, with a representative distribution of demographics and occupations. Networks of contacts are established to simulate potentially infectious interactions though occupation, household, and random interactions

**Interventions:** we simulate standard Covid-19 vaccination, versus delayed-second-dose vaccination prioritizing first dose. Sensitivity analyses include first-dose vaccine efficacy of 70%, 80% and 90% after day 12 post-vaccination; vaccination rate of 0.1%, 0.3%, and 1% of population per day; assuming the vaccine prevents only symptoms but not asymptomatic spread; and an alternative vaccination strategy that implements delayed-second-dose only for those under 65 years of age.

**Main outcome measures:** cumulative Covid-19 mortality over 180 days, cumulative infections and hospitalizations.

**Results:** Over all simulation replications, the median cumulative mortality per 100,000 for standard versus delayed second dose was 226 vs 179; 233 vs 207; and 235 vs 236; for 90%, 80% and 70% first-dose efficacy, respectively. The delayed-second-dose strategy was optimal for vaccine efficacies at or above 80%, and vaccination rates at or below 0.3% population per day, both under sterilizing and non-sterilizing vaccine assumptions, resulting in absolute cumulative mortality reductions between 26 and 47 per 100,000. The delayed-second-dose for those under 65 performed consistently well under all vaccination rates tested.

**Conclusions:** A delayed-second-dose vaccination strategy, at least for those under 65, could result in reduced cumulative mortality under certain conditions.

## Introduction

The global public health response to the Coronavirus 2019 (COVID-19) pandemic has resulted in the massive investment of resources into the production of an effective vaccine [1]. This unparalleled approach has led to the development of multiple effective vaccines in record time. The first two vaccines to be approved in the United States (US), BNT162b2 (sponsor: Pfizer/BioNTech) and mRNA-1273 (sponsor: Moderna) both use a messenger ribonucleic acid (mRNA) encoding the SARS CoV-2 spike protein. These mRNA vaccines are both two-dose regimens, with the second dose administered 21 or 28 days following the initial dose [2, 3]. An additional viral vector vaccine based on ChAdOx1 (sponsor: AstraZeneca) also received approval for use in most of Europe and the UK [4].

In the face of persistently high rates of COVID-19 infection, the emergence of new strains such as B.1.1.7 (UK), B.135 in South Africa and P.1 in Brazil, and an ever-increasing death toll, there is mounting pressure to achieve population-wide immunity as quickly as possible [5-9]. However, the ability to achieve sufficient levels of immunity has been hampered by a vaccine supply and vaccination logistics that is far outstripped by demand.

Multiple public health authorities have proposed prioritizing single-dose vaccination for as many people as possible, even if this means delaying a second dose beyond the studied 21- or 28-day time frame[10-12]. The justification for this relies on the assumption that meaningful protection against COVID-19 can be achieved after a single vaccine dose, a point which is the subject of intense debate. Those taking a conservative interpretation of available data argue that a delayed second dose regimen was not explicitly studied in clinical trials, nor was the possibility of asymptomatic infectious spread, therefore, public health agencies should practice only the regimens explicitly studied to be certain of the results they will achieve. Others more willing to extrapolate from clinical trial results based on prior immunological research argue that there is probably meaningful protection against COVID-19 after one vaccine dose. Recent calculations using clinical trial data have estimated first dose vaccine efficacy for the Pfizer and Moderna vaccines to be 92.6% and 92.1%, respectively [13]. This has led some authors to suggest a delayed-second-dose strategy to reduce the cumulative mortality [10, 13]. However, the risk of infection is dependent on complex network dynamics, and the case fatality rate can be up to two orders of magnitude higher for different demographic groups[14]. Estimating the impact of different vaccination strategies requires the use of methods that can take these non-linear effects into consideration.

Therefore, to investigate we used agent-based modeling (ABM) to measure the relative impact of delaying second dose vaccine policies on infections, hospitalizations and mortality compared to the current on-schedule two dose regimen. To account for uncertainty, we used sensitivity analysis and examined multiple different scenarios such as whether the vaccine offers sterilizing, versus only symptomatic, immunity. We also examined a novel dosing strategy in which a delayed second dose regimen is used for those younger than 65 years old, but the on-schedule two dose is used for those older.

## Methods

We extended an open-source ABM from the literature to model the impact of the delayed second dose versus standard dosing vaccination strategies on COVID-19 infections, hospitalizations and deaths in a population with 100,000 agents over a time period of six months [15]. The results were aggregated over 10 runs of the simulation. The original open-source ABM was limited to modeling COVID-19 spread [15]. Our extension improved the processing speed by using matrix computation and added the possibility of implementing different vaccination policies. We used Python 3.7, the full list of packages can be found in the supplemental material.

In the model, agents interact with each other in three types of networks: an occupation network, a family network, and a random encounter network. Each encounter between an infectious and a susceptible agent has a probability of infection transmission. Once infected, agents have a certain probability of being asymptomatic; if they are symptomatic, they have a pre-symptomatic period, followed by a probability distribution of symptom severity and a subsequent probability distribution of death. Our assumptions regarding disease progression, transmission characteristics, and family, occupation, and random network interactions are the same as in the original ABM and are available in our supplemental materials [15].

In addition, we explicitly model the confirmation of infections with PCR testing, quarantining of known infected subjects with imperfect compliance over time. We report results on relevant outcomes (deaths, cumulative infections, and fraction immune) averaged over 10 replications of our ABM simulation. To simulate a natural pattern of infection at the point vaccinations are started, we initiate our simulation with 10 agents infected and run the simulation for 20 days before starting vaccinations, which corresponds to a cumulative infection rate of 1%, similar to the one in the US, UK and most of Europe when vaccinations were started. We then run the simulation for a total of 180 days, using discrete time by day. In all our vaccination strategies, we start administering vaccines based on age, starting with those over 75, then those over 65, and so on. For further information regarding the ABM and the exact vaccine prioritization under each strategy considered, please see the supplemental section.

### Vaccine and infection characteristics

We conducted four analyses to derive insights about four different model parameter variations. In all our analyses, vaccines are administered in an age-prioritized fashion, with the oldest individuals receiving their vaccines first, regardless of the vaccine regimen examined. For comparisons of our vaccine regimens studied see the supplemental section. Sections 1–3 assume that COVID-19 vaccines prevent both symptoms of infection and transmission of virus (sterilizing vaccine), while Section 4 examines the possibility that vaccines prevent only symptoms but not asymptomatic infection and spread (non-sterilizing vaccine). All simulations assume a VE of two vaccine doses of 95%.

Our first analysis sought to understand potential risks or benefits of delayed second dose versus standard dosing strategies under varying estimates of single-dose efficacy. In this analysis, we examined outcomes of deaths, hospitalizations, and infections. To model single-dose efficacy, we assumed no protection against COVID-19 infection for the first 12 days after the initial dose, and thereafter a protection of 70%, 80% or 90% which persists for the remainder of our 180-day simulation. We selected these estimates based on examination of the BNT162b2 Pfizer trial results, which showed that between days 1 to 11, the number of cases was similar between the vaccinated and unvaccinated groups. Between days 12 and 21, there were 4 and 30 infections in the treatment and control arms, respectively. This suggests a vaccine efficacy from a single dose of 87%. Assuming cases in the vaccine and control groups follow two different Poisson random distributions, based on the trial data the 95% confidence interval for the rate ratio between them (which corresponds to the vaccine effectiveness) is 66% to 96%. We set the vaccination rate to doses per day of 0.3% of the population.

Our second analysis examined the effect of varying vaccination rates on total deaths using the same two dosing regimens as our first analysis, and a fixed single-dose efficacy estimate of 80%, which we considered relatively conservative given our point estimate of 87%. For this analysis, we used vaccination rates of 0.1%, 0.3% and 1% doses administered per person per day. The vaccination rate in the US was around 0.1% per day as of February 2021, according to data from the CDC [16]. The vaccination rate of 0.33% per day approximately corresponds to the goal laid out by President Biden to vaccinate 100 million individuals in 100 days, while 1% per day represents an ambitious future possibility.

Our third analysis examined the utility of an additional age-split vaccination strategy, across the vaccination rates used in our second analysis, for preventing death. This additional vaccination strategy proposed to use a delayed second dose strategy in those under 65 years old, and a standard dose regimen for those 65 and above. We propose this strategy based on the fact that elderly individuals have the highest mortality risk, and therefore, providing them with maximal vaccine protection is likely to avert the most deaths.

We also performed a sensitivity analysis to consider the possibility that the vaccine prevents only symptomatic disease but not asymptomatic infection and spread. In this analysis, we replicated the methodology of our first analysis, using different single-dose efficacy rates and fixed administration rate of 0.3% of the population per day, but annulled the assumption that the vaccine prevents asymptomatic spread.

We display our results using time-series line plots over our 180-day model period, with a central line corresponding to the median value of our 10 runs for a given day, and a shaded band corresponding to the 25-75% percentile of values for a given day across all runs.

## Results

We present our results in four sections. Section 1 examines the effect of different single-dose efficacy estimates on outcomes. Section 2 describes the effects of different rates of vaccination. Section 3 examines the effect of a hypothetical vaccine regimen in which second doses are delayed only for those under age 65. Section 4 replicates the analysis of Section 1 with the modification that the vaccine only prevents symptoms, but not asymptomatic spread.

### Section 1: Effect of standard vs delayed second dose regimens using various efficacy estimates, with intermediate vaccination rate

Figure 1 presents the results for cumulative mortality comparing the standard vaccination strategy and a delayed-second-dose vaccination strategy under three different values of the first-dose efficacy: 70%, 80% and 90%, all with intermediate vaccination rate of 0.3%. Total mortality per 100,000 for standard versus delayed second dose is 226 vs 179; 233 vs 207; and 235 vs 236; for 90%, 80% and 70% first-dose efficacy, respectively. These results suggest that higher first-dose efficacy estimates favor delaying the second dose, and that for a first-dose efficacy of 70%, there seems to be no meaningful difference between the standard and delayed-second-dose strategy.

**Figure 1.**
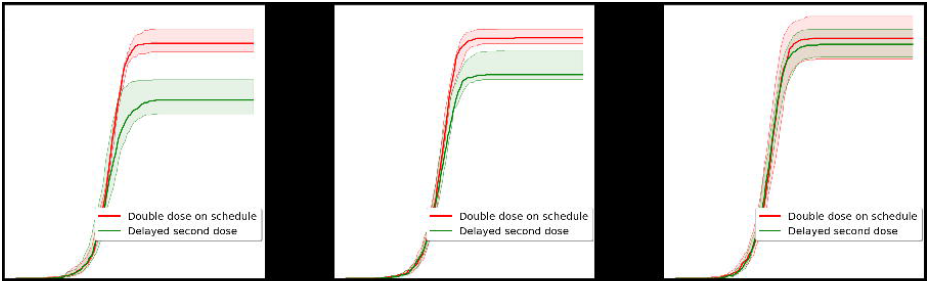
Comparison of cumulative mortality for delayed-second-dose versus standard vaccination strategy under three different first-dose effectiveness assumptions. Results are shown for a vaccination rate of 0.3% of the population per day. The total cumulative mortality on day 180 is lower for the delayed-second-dose scenario under the assumption that the first-dose effectiveness is at least 80%.

Figure 2 presents the total number of infections and the number of hospitalizations for the same efficacy estimates and vaccination regimens. Cumulative number of infections per 100,000 for standard versus delayed second dose is 69,577 vs 64,220, 69,350 vs 64,859, and 69,670 vs 65,891; for 90%, 80% and 70% first-dose efficacy, respectively. Thus, the number of cumulative infections is similar between the two strategies in the three scenarios studied.

**Figure 2.**
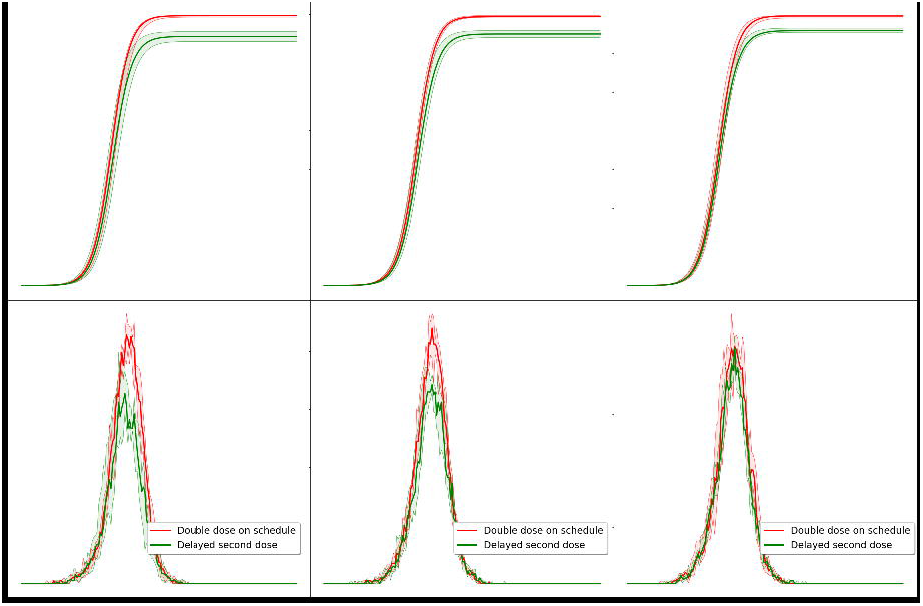
Results are shown assuming a vaccination rate of 0.3% of the population per day. The cumulative number of infected on day 180 is lower for the delayed-second-dose approach. The peak hospital census becomes similar for both approaches as the efficacy of the first-dose vaccine is reduced.

### Section 2: Effect of standard vs delayed second dose regimens using various vaccination rates, with single-dose vaccine efficacy held constant at 80%

Figure 3 presents the cumulative mortality in three different vaccination rate scenarios in which 0.1%, 0.3%, or 1% of the population is vaccinated per day. These results suggest that at a single-dose vaccine efficacy estimate of 80%, population mortality is lower when the second vaccine dose is delayed, except in scenarios of extremely high vaccination rates, more than current practices.

**Figure 3.**
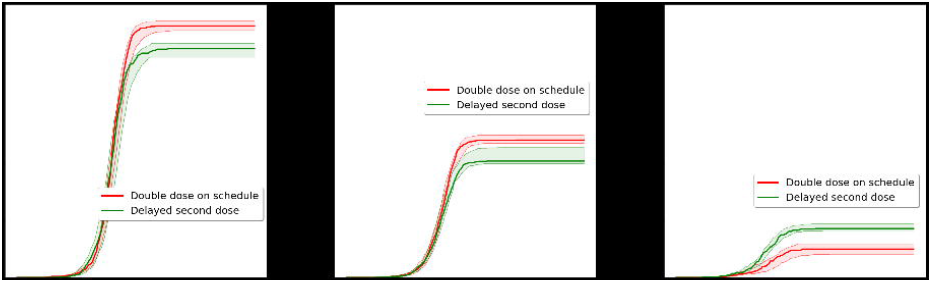
Comparison of cumulative mortality for delayed-second dose versus standard vaccination strategy under three different vaccination rate assumptions. The impact of vaccination rate on the comparative effectiveness of double dose on schedule and delayed second dose strategies in reducing cumulative mortality.

As mentioned before, this corresponds roughly to the current vaccination administration rate in the US and Europe (0.1% population per day); the current vaccine production rate (0.3% population per day); and the estimate of increased vaccine production rate once more vaccines are available (1% population per day). The number of total deaths is lower for higher vaccination rates, with the optimal strategy switching at a value between 0.3% and 1%. Total estimated mortality per 100,000 for delayed vs standard second dose is 402 vs 442, 204 vs 241, and 85 vs 50, for vaccination rates of 0.1%, 0.3% and 1%, respectively.

This suggests the delayed-second-dose strategy is optimal for vaccination rates at or lower than 0.3% population per day if the VE from one dose is 80%.

### Section 3: Effects of additional age-split dosing strategy at different vaccination rates, single dose vaccine efficacy held constant at 80%

Figure 4 explores the effect on cumulative mortality of an additional vaccination strategy that prioritizes second doses for those older than 65 years, across three different vaccination rates. The total number of deaths is lower for higher vaccination rates, as expected. This strategy, that we call “delay second dose except for 65+”, has lower cumulative mortality than the standard strategy for low and medium vaccination rates (0.1% and 0.3%), and a lower mortality than the delayed second dose strategy for high vaccination rates (1%).

**Figure 4.**
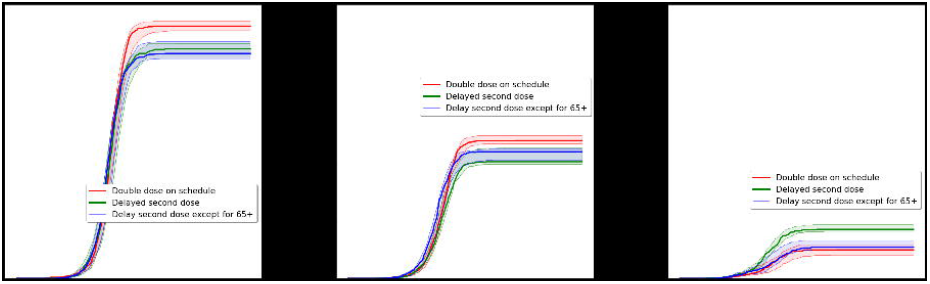
Comparison of cumulative mortality for three different vaccination strategies (delayed-second-dose, versus standard vaccination, versus delayed-second-dose except for 65+) under three different vaccination rate assumptions. The “delayed second dose except for 65+” (blue line) strategy seems optimal or close to optimal under all three assumptions, making it a safe choice in the face of an uncertain future vaccination rate.

The cumulative mortality rate for delayed vs standard vs “delayed except for 65+” strategies is 402 vs 442 vs 394; 204 vs 241 vs 222; and 86 vs 50 vs 55. This suggests that the “delayed second dose except for 65+” strategy is optimal or close to optimal assuming a conservative first-dose efficacy of 80%, and for vaccination rates at or below 0.3% population per day.

Figure 5 presents similar results to Figure 1, but this time under the assumption that the vaccine only prevents symptoms, but not infection spread. Under this assumption, with a vaccination rate of 0.3% population per day, the estimated cumulative mortality for delayed versus standard second dose are 179 vs 226; 207 vs 233; and 235 vs 236; for a first-dose effectiveness of 90%, 80% and 70%, respectively. The delayed second dose strategy seems optimal or close to optimal for a one-dose vaccine efficacy of at least 70%.

**Figure 5.**
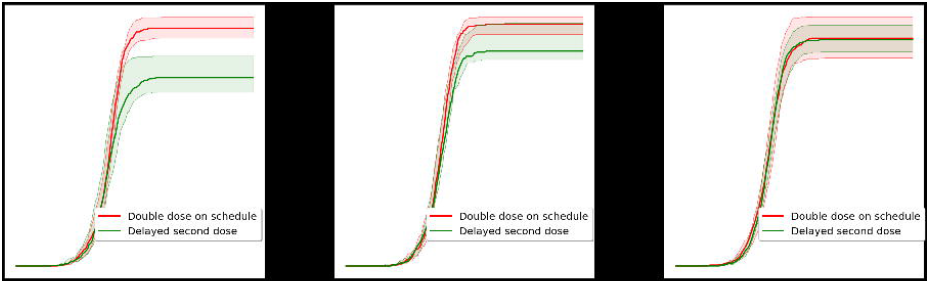
Cumulative mortality for delayed-second-dose versus standard dosing under a non-sterilizing vaccine assumption. The results are similar to those under a sterilizing vaccine assumption: the delayed second dose strategy seems optimal for a first-dose efficacy of at least 80%.

## Discussion

Our study compared two COVID-19 vaccine strategies that delayed the second dose to the on-schedule two dose strategy that is currently being used in the US. The results suggest under specific conditions a decrease in cumulative mortality, infections, and hospitalizations can be achieved when the second vaccine dose is delayed. This was most significant in the group where the vaccine was delayed in those below 65 years of age, but not in those older. The conditions where these benefits were observed included the first dose vaccine efficacy being above 70% and vaccination rates remaining below 1% of the population per day. These two conditions seem reasonable based on available data [16, 17]. The timeframe of 180 days used in our study was thought to be important to policy makers who face the immediate challenge of increasing population immunity with the current vaccine production rates [18].

Our findings suggest vaccination rate is an important factor in choosing a strategy. A delay in second dose strategy either in those below 65 years old or the entire population did not show a cumulative mortality benefit compared to an on-schedule two dose regimen when the vaccination rate is 1% of the population or above. At very low vaccination rates the differences in delay strategy were not observed but favored delays in 65-year-old and younger when rates were 0.3% of the population per day, approximately the current US rate [16]. Our findings also suggest that changes in cumulative mortality are larger than the corresponding decrease in the number of infections. For example, the relative reduction in the cumulative number of infections for a vaccination rate of 0.3% and a first-dose efficacy is around 6%, whereas the reduction is 11% for mortality.

These results may be broadly informative for COVID-19 strategy. In the US, the national strategy continues to be a strict two-dose schedule for either BNT162b2 or mRNA-1273 vaccine [19]. But, while our vaccination administration rates are based on current US capabilities, the intuition derived from simulation of multiple parameters may be useful for policymakers elsewhere. With the continued large death toll from COVID-19 globally and reports of mutant strains there is increasing urgency to vaccinate the population rapidly [7, 20]. The multiple vaccines in stage III trials bring promise to increasing availability and therefore vaccination rates, but the timeline could be at least 6 months [21]. The strategy of delaying the second dose has been an active discussion but lacked empirical research to understand its implications.

The primary strength of our study is the use of agent-based modeling to forecast the effects of different vaccine strategies across a timeframe that is useful to decision makers, while capturing the complexity of human interactions, which are critical in COVID-19 transmission. Additionally, our results are strengthened using 100,000 agents with age-based demographics reflective of a sample population in the US, simulated over various human interaction networks reflecting real-world behavior, and for a duration of 180 days.

Our model estimates that, without any intervention, the infection spreads to saturation within 180 days. This may be a pessimistic estimate if more aggressive infection containment measures are put in place, such as stricter lockdowns, or higher compliance with public health recommendations. However, the key parameter is the relationship between vaccination rate and infection spread and this would likely not affect the relative effectiveness of different vaccination strategies.

We identified three primary limitations of our study design: (1) the paucity of clinical trial data regarding first dose vaccine efficacy, (2) information on immune decay in first dose only recipients, and (3) whether the vaccine is sterilizing or not. Our study did not measure the impact of COVID-19 mutant strains and various infectivity rate, or differences in behavior geographically, or the impact of other preventative measures such as digital exposure notification or testing availability and turn-around-times that are variable state-to-state[22]. These are limitations but would not likely change the relative differences measured between strategies.

In the BNT162b2 and mRNA-1273 vaccine trials, the single dose vaccine efficacy was initially reported to be 52% and 80%, respectively. This was estimated in the small subset of subjects who did not receive the second dose during the trial [2, 3, 23]. Since these were not defined study sub-groups the advantages of randomization in preventing bias cannot be assumed to hold true, and in fact unknown bias in these individuals is likely. This limitation cannot be overcome using simulation modeling. However, we believe reasonable estimates can be made using the data available. The 52% vaccine efficacy in the Pfizer study was attributed to inclusion of the first 12 days after vaccination in their estimate. Using the 12 days underestimates the true vaccine efficacy because sufficient time to develop immunity had not occurred. This is well established in vaccine and immunity literature and holds true regardless of vaccine type. In our study we re-estimated the BNT162b2 vaccine efficacy to be approximately 87%, and this fits with the mRNA-1273 reported estimate. Additionally, the use of sensitivity analysis for a range of first dose vaccine efficacies of 70-90% reduces this limitation in our study design.

To address immune decay, we analyzed existing literature on vaccines and specifically mRNA/DNA vaccine platforms. In particular, recipients of DNA vaccines against MERS and SARS had waning immunity, generally after 6 months. Clinical trials of a flu H7N9 mRNA vaccine showed detectable antibody titers in participants after 6 months suggesting that a time frame of delay of 6 months from first dose could maintain functional immunity[24]. In preclinical data in non-human primates, a single dose of the Ad26.COV2.S vaccine resulted in stable antibody responses up to 14 weeks[25], but longer time frames have not been reported. In fact, a delay of the second dose from 4 to 8 weeks resulted in higher neutralizing antibody titers[26]. There is no evidence to date that a 6-month dose delay would impair the efficacy of the second vaccine.

To understand the impact of whether the vaccine is sterilizing (prevents transmission and serious symptoms) or non-sterilizing (only prevents serious symptoms, including death) on the outcomes between the vaccine strategy groups, we modeled both scenarios. In either case, the differences between vaccine strategy groups did not meaningfully changed. Although lack of data about the sterilizing properties of either the BNT162b2 or mRNA-1273 vaccine is a limitation, our analysis of both scenarios was a strength of our study design.

To date, our study is the first to analyze the impact of delaying a second dose for the BNT162b2 and mRNA-1273 vaccines under conditions we believe are necessary for decision makers considering second dose delay strategies. Moreover, our study is the first to look at varying the second dose delay in only those younger than 65 years old. There is a pre-print study also analyzing this question using an agent-based model, but their design used fixed delay periods, a shorter time horizon, and did not perform sensitivity analysis on various first dose vaccine efficacies or non-sterilizing vaccine effectiveness, but it suggested that if first dose vaccine efficacy is 80% a delayed second dose strategy is optimal [27]. Another recent study, currently under review at The Lancet, randomized individuals to a delayed dose of 12 weeks or longer of ChAdOx1 nCoV-19 (AstraZeneca) vaccine[4]. The AstraZeneca vaccine is adenovirus-based and has a lower overall effectiveness, versus Pfizer or Moderna’s mRNA vaccine, therefore comparisons should be done cautiously. The results showed AstraZeneca’s single dose vaccine efficacy to be 76% after 21 days and showed negligible immune decay over 3 months. Interestingly, their study also found that delaying the second dose boosted the second dose efficacy compared to the typical two dose schedule of 22 days apart [4].

The COVID-19 pandemic continues to take thousands of lives daily. The promise of vaccines mitigating the pandemic has been overshadowed by disappointment about the vaccine roll out. Decision makers are looking for short-term solutions until more vaccines are available in the next 6 months. The supply and logistics of delivering a regimented two dose schedule of the BNT162b2 and mRNA-1273 vaccines have proven challenging. Delaying the second dose of either of these vaccines has been an appealing strategy because it would significantly increase vaccine availability and reduce the logistics of a strict two dose schedule. Hesitation regarding delaying a second dose is understandable given limitations of any study design that is not a randomized trial. However, our agent-based model can provide estimates of relative differences between these strategies that can be helpful in making policy decisions. The risks associated with delaying a second dose could also be mitigated by selectively doing so in those younger than 65, who have approximately a 10 times lower risk of mortality than those 75 or above and likely more robust immune response to single dose vaccination. Importantly, our results suggest this may also be the optimal strategy to prevent deaths. This could provide reassurance in those who have hesitancy about a delay strategy. Decision makers will need to weigh the risks, mostly from uncertainty of such strategies not evaluated in randomized trials, to possible benefits of preventing COVID-19 deaths.

## Data Availability

All data and code used will be available in our GitHub repository.

https://github.com/ayushchopra96/deepabm-covid.git&#8203;

## Notes

### Competing Interest Statement

The authors have declared no competing interest.

### Funding Statement

All research was supported with volunteer time and internal funds from our respective research institutions: MIT, ASU and Mayo Clinic. No third-party funding was received for this work.

### Author Declarations

Mayo Clinic Institutional Review Board

