## Supplemental files for "The Public Health Impact of Delaying a Second Dose of the BNT162b2 or mRNA-1273 COVID-19 Vaccine"

**APPENDIX: Supplemental Material on Details of the Agent-Based Model (ABM)**

Our agent-based simulation model and point estimates for parameters are based on the model given in~\citep{Hinch2020.09.16.20195925}.   We simulate a total of 100,000 agents, each of whom interact with a subset of other agents. These interactions serve as the route of transmission of the disease. We consider three types of agent interactions, namely, household interactions, occupational interactions and random interactions. These are modeled using three respective networks, which are described in detail below. In addition to these interactions, agents' choices in terms of getting tested and quarantining also influences infection spread.

To analyze the impact of different vaccination strategies, we extended the model provided by ~\citep{Hinch2020.09.16.20195925} to implement the prioritization logic with which vaccination takes place. In particular, our model includes a meta-agent that determines the vaccination strategy, which is the final factor that influences infection spread. We have also extended the simulation model to represent the building of immunity in vaccinated individuals with their first and second doses. All of the relevant parameters are sourced from published trial data and references, as explained in the main body of the paper.

In particular, we model a single type of vaccine that requires a two-dose regimen, where the second dose may be taken any time 21 days after the first dose. All other things being equal, under each vaccination strategy, we assume that an older individual is prioritized over a younger individual for vaccinations. We consider three vaccination strategies, all defined by the order in which first-dose eligible and second-dose eligible individuals of different ages are prioritized.

To demonstrate the three vaccination strategies clearly, we offer the following example of six hypothetical individuals:

Adam: First dose eligible 78 yr old

Betty: Second dose eligible 78 yr old

Charlie: First dose eligible 68 year old

David: Second dose eligible 68 year old

Eleanor:First dose eligible 40 year old

Frank: Second dose eligible 40 year old

The last column of the table below indicates the order in which these individuals would be vaccinated under each vaccination strategy.

**Table A1. Definitions of the three vaccination strategies with a prioritization example**

| **Name of vaccination strategy** | **Description** | **Priority list under strategy will be** |
| --- | --- | --- |
| Standard strategy  (Double dose on schedule) | Age prioritized vaccination with second dose at 21 days | Betty, David, Frank, Adam, Charlie, Eleanor |
| Delayed second dose | Age prioritized vaccination prioritizing vaccination of first-dose eligible individuals | Adam, Charlie, Eleanor, Betty, David, Frank |
| Delayed second dose except for 65+ | Age prioritized vaccination with first dose prioritization, except for the elderly (above 65), who receives a second dose on schedule | Adam, Betty, Charlie, David, Eleanor, Frank |

Under each vaccination strategy, we run the 10 replications of the simulation for 180 days, and observe the number of infections, hospitalizations and deaths over time for each simulation replication. We initialize the simulations with 10 infected agents and start vaccinations when the number of infected agents reach 1% of the population, which happens around simulated day 20.

We plot the median as well as 25th and 75th percentiles of those 10 runs for each outcome of interest under the vaccination strategies to compare and contrast them with respect to their effectiveness in reducing the number of cumulative deaths as well as cumulative infections and the number of hospitalizations over time. In the following sections, we explain the different components of our ABM.

**State space:** At any given time, each agent occupies a state that consists of static components (i.e., context) as well as dynamic components. The static components (i.e., context) are attributes of the agent that do not change over the course of the simulation. We keep the following static components for each agent:

1. Age
2. Household
3. Occupation
4. Number of random interactions in a day
5. App user status (for digital exposure notification)

The dynamic components, on the other hand, are attributes that change over time, and are used to drive the various events (like transmission instances) in the simulation.

1. Disease stage that the agent currently occupies (susceptible, infected asymptomatic, presymptomatic mild, presymptomatic severe, mild symptomatic, severe symptomatic, hospitalized, critical & ICU, recovered, dead)
2. Disease stage transition times (randomly generated for the agent from the assumed distributions)
3. Test dates
4. Test results (positive, negative; randomly generated with the assumed test sensitivity and specificity)
5. Quarantine status
6. Vaccination status

**Action space:** At each step (i.e., day), if the agents have not already tested positive or are not in a hospitalized, recovered or dead state, they have an option to get tested. For positive test results, the agents who are not in a hospitalized or recovered or dead state have an option to quarantine themselves. Furthermore, agents who are currently in the quarantined state have the option to break quarantine (with a small probability). We detail these actions/interventions in the next section.

**Networks:** Each agent is part of three networks, which are constructed based on the method described in~\citep{Hinch2020.09.16.20195925}. We summarize the network construction process below, for each of the three networks.

1. *Household network*: All agents within the same household are connected to each other.
2. *Occupation network:* For each step, a Watts-Strogatz network is constructed for agents having the same occupation, where the parameters of the network are occupation-dependent.
3. *Random network:* Using each agent's number of random interactions, a random network that includes all agents is constructed.

**Transition dynamics:** The transition dynamics in our ABM involves specifying the temporal evolution of the state variables of all the agents in the systems, given the actions taken by the agents or the meta-agent. Of these state variables, the test, quarantine and vaccination state variables have a relatively straightforward evolution given the agents' actions for the first two and the meta-agent's action for the last. The evolution of the agent's stage is more complicated and is based on the model given in~\citep{Hinch2020.09.16.20195925}. The stage dynamics, which are modified SEIR dynamics at the agent level, can be divided into two parts: (i) infection dynamics, and (ii) disease dynamics.

**(i) *Infection dynamics:*** This part involves the state transition from the susceptible to either asymptomatic or pre-symptomatic infection states. This step involves aggregating the effect of interacting with all other agents taking into account the quarantine and vaccination states of the various agents. This is a Markovian transition where at each step there is a probability of transitioning from a non-infected susceptible state to an infected state. The infectiousness of each interaction is determined based on the ages, disease stage, quarantine and vaccination status of the agents, and the network that supports this interaction. The probability of infection is computed by aggregating the infectiousness factor from each interaction.

**(ii) *Disease dynamics:*** This part involves transition between the various infected disease stages ultimately leading to either a recovered or a death absorbing state. These transitions are semi-Markov, where the transition or sojourn times are drawn from continuous time distributions with disease stage and age-dependent parameters.

**Modeling Interventions:** Our ABM supports several interventions for mitigating the transmission of COVID-19. In this section, we include some of the baseline assumptions used for our experiments.

***Quarantine Behavior:***  An agent who tests positive quarantines himself for a period of 14 days following the test result. To model waning compliance behavior, we also define a per-day quarantine drop-out probability of 2% per day.

***Testing:*** Symptomatic agents are assumed to opt for tests. In the baseline case, agents take an RT-PCR test and get results in 2, 3, or 4 days. We assume that the COVID-19 test used has a specificity of 0.90 and sensitivity of 0.99, which is consistent with the state-of-the-art tests used today.

***Vaccine effect on immunity***: When an agent receives a vaccine dose, they have a certain probability of becoming immune. This probability is dependent on whether it’s their first dose or their second dose. Twelve days after receiving the first dose of the vaccine, agents have a 70%, 80% or 90% probability of becoming immune (depending on the scenario). After receiving the second dose, agents have a 95% probability of becoming immune. In most scenarios, immune agents cannot become infected, and thus cannot transmit the disease. In the last scenario tested (that we named “non-sterilizing vaccine”), immune patients are only “immune” in the sense that they cannot become symptomatic, but they can still become asymptomatically infected and transmit the disease with the same probability as a non-vaccinated asymptomatic patient.
